# Different pathways toward net-zero emissions imply diverging health impacts: a health impact assessment study for France

**DOI:** 10.1101/2023.10.03.23296478

**Authors:** Léo Moutet, Aurélien Bigo, Philippe Quirion, Laura Temime, Kévin Jean

## Abstract

**Background:** In the transport sector, efforts to achieve carbon neutrality may generate public health cobenefits by promoting physical activity.

**Objective:** This study aims to quantify the health impacts related to active transportation based on four different scenarios leading France toward carbon neutrality in 2050.

**Methods:** The French Agency for Ecological Transition developed four consistent and contrasting scenarios (S1 to S4) achieving carbon neutrality by 2050 as well as a business-as-usual (BAU) scenario that extends our current lifestyles until 2050, without reaching net-zero. For each of these *Transitions2050* scenarios, we distributed the mobility demand for walking, cycling and e-cycling across age groups. Relying on the health impact assessment framework, we quantified the impacts of the corresponding physical activity on all-cause mortality. The impact of each of the carbon neutrality scenarios was determined by comparison with estimates from the BAU scenario.

**Results:** In S1 and S2 scenarios, volumes of active transportation are projected to increase to fulfil the World Health Organisations recommendations by 2050, while they increase slightly in S3 and decrease in S4. S2 scenario reaches the highest levels of health cobenefits, with 494,000 deaths prevented between 2021 and 2050. This would translate into a life expectancy gain of 3.0 months for the general population in 2050, mainly driven by e-bikes. S1 would provide smaller but important health benefits, while these benefits would be modest for S3. On the contrary, S4 implies 52,000 additional deaths as compared to the BAU scenario, and a loss of 0.2 month in life expectancy.

**Discussion:** Different ways to decarbonize mobility in a net-zero perspective may achieve very contrasting public health cobenefits. This study illustrates how the public health dimension may provide a relevant insight in choices of collective transformation toward net-zero societies.

## Introduction

The Paris agreement, adopted in 2015, establish the objective of constraining the rise in average global temperature to 2°C above preindustrial levels (UNFCCC (United nations Framework Convention on Climate Change), 21st session, 2015). To achieve this, a substantial reduction of 43% in greenhouse gas (GHG) emissions by 2030 is necessary (IPCC (Intergovernmental Panel on Climate Change), 2023). The sixth report from the Intergovernmental Panel on Climate Change (IPCC) highlights the importance of numerous actions that effectively reduce our carbon footprint while concurrently promoting human health. These health co-benefits manifest across several dimensions such as diet, transport, energy supply or green and blue infrastructures (IPCC (Intergovernmental Panel on Climate Change), 2023).

The transport sector accounted for approximately 15% of anthropogenic GHG globally in 2019 (IPCC (Intergovernmental Panel on Climate Change), 2023). In France, it contributes up to 31% of GHG emissions (HCC (The High Council on Climate), 2021). Amongst the many existing levers to limit these emissions (electrification of the energy supply, increased vehicle occupancy, lower transport demand…), some have the potential to yield health co-benefits. Among those, shifting to active modes of transportation offers the additional advantage of mainly relying on local decisions such as reallocation of existing infrastructures and land use planning (Stappers et al., 2023).

Active transportation encompasses essential daily activities (e.g., commuting to work, visiting the doctor, shopping) as well as leisure activities (e.g., cultural outings, sports) and has indeed been identified as a lever for achieving socio-economic, air quality, and public health co-benefits (IPCC (Intergovernmental Panel on Climate Change), 2023). However, physical activity levels worldwide have failed to meet international recommendations, affecting both adolescents and adults (Guthold et al., 2018). In high-income countries, 9.3% of overall mortality can be attributed to physical inactivity (Katzmarzyk et al., 2021). Insufficient physical activity also incurs substantial monetary costs for public and private healthcare systems and households while leading to decreased productivity (Ding et al., 2016). A 2017 study focusing on urban populations estimated that meeting international guidelines on physical activity, air pollution, noise, heat, and access to green spaces could prevent 20% of deaths. Among these factors, physical activity yielded the largest proportion of preventable deaths (Mueller et al., 2017).

The health benefits associated with active transportation may be substantiated by various health impact assessment methods that consider diverse parameters, regardless of the geographical context (Mueller et al., 2015). Institutions and organisations have devised prospective pathways outlining the necessary transformations required for the societies to attain the objectives of the Paris agreement. Numerous studies quantify the health impacts of active commuting in prospective scenarios, with a majority conducted in Europe, North America or Oceania, primarily focusing on urban areas (Mueller et al., 2015; Rojas-Rueda et al., 2016). Nevertheless, to the best of our knowledge, only one study has been conducted at the national level in France, quantifying the impacts of a single scenario compared to the baseline levels of 2021 (Barban et al., 2022).

In this study, we quantified the health impacts related to physical activity through active transportation based on four consistent and contrasting scenarios leading France toward carbon neutrality in 2050.

## Methods

### The ADEME Transition scenarios

In 2021, ADEME (The French Agency for Ecological Transition) released the outcomes of a prospective project *Transitions2050*, describing four scenarios (S1 to S4). Each of these scenarios is designed to achieve carbon neutrality by 2050 in metropolitan France following distinct trajectories corresponding to different societal choices (Figure 1) (The French Agency for Ecological Transition, n.d.). Additionally, a business-as-usual scenario (BAU) was formulated, representing a projection of our current lifestyle trends until 2050, without achieving carbon neutrality. It partly corresponds to an extension of certain past trends and partly to evolutions already underway.

**Figure 1.**
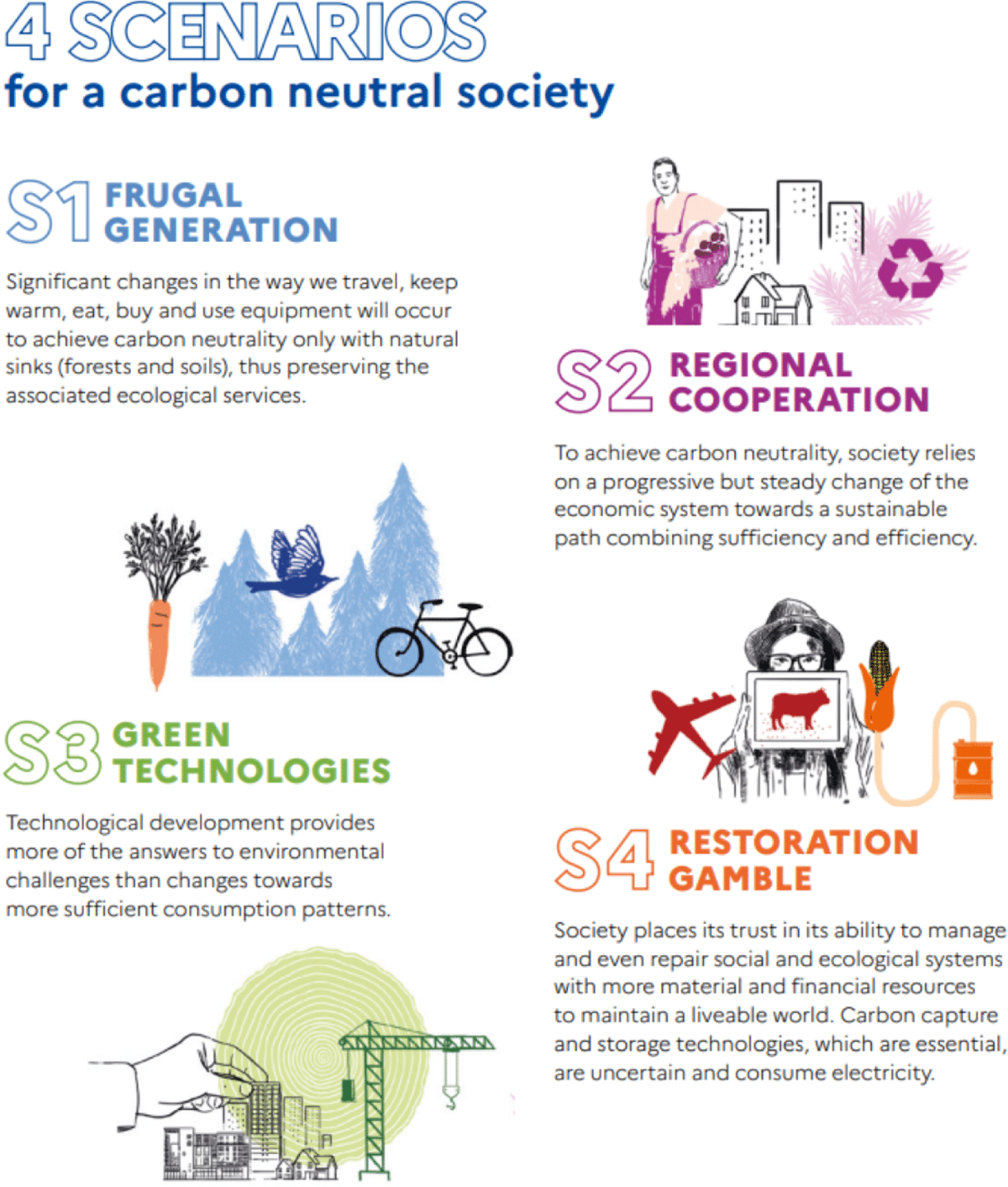
Summary of ADEME’s prospective carbon neutral scenarios. Credit: @ADEME - <u>Transition(s) 2050</u> - Illustrations: S. Kiehl

Each *Transitions2050* scenario describes the evolution of the main energy consumption drivers over time, especially the demand for transportation for various modes of transportation (Table 1 & Table S1). Within these estimates, three active modes are considered: cycling, E-cycling and walking, without accounting for recreational activity. Note that the estimates for walking encompass distances covered while transitioning to or from another mode of transport, especially public transportation (e.g train or bus). For each active travel mode, the scenarios express the yearly national demand in terms of Giga passenger-kilometres (Gpkm) at five-year intervals over the period 2015-2050. For this study, passenger-kilometres values were linearly interpolated for each year within the intervals.

**Table 1.**
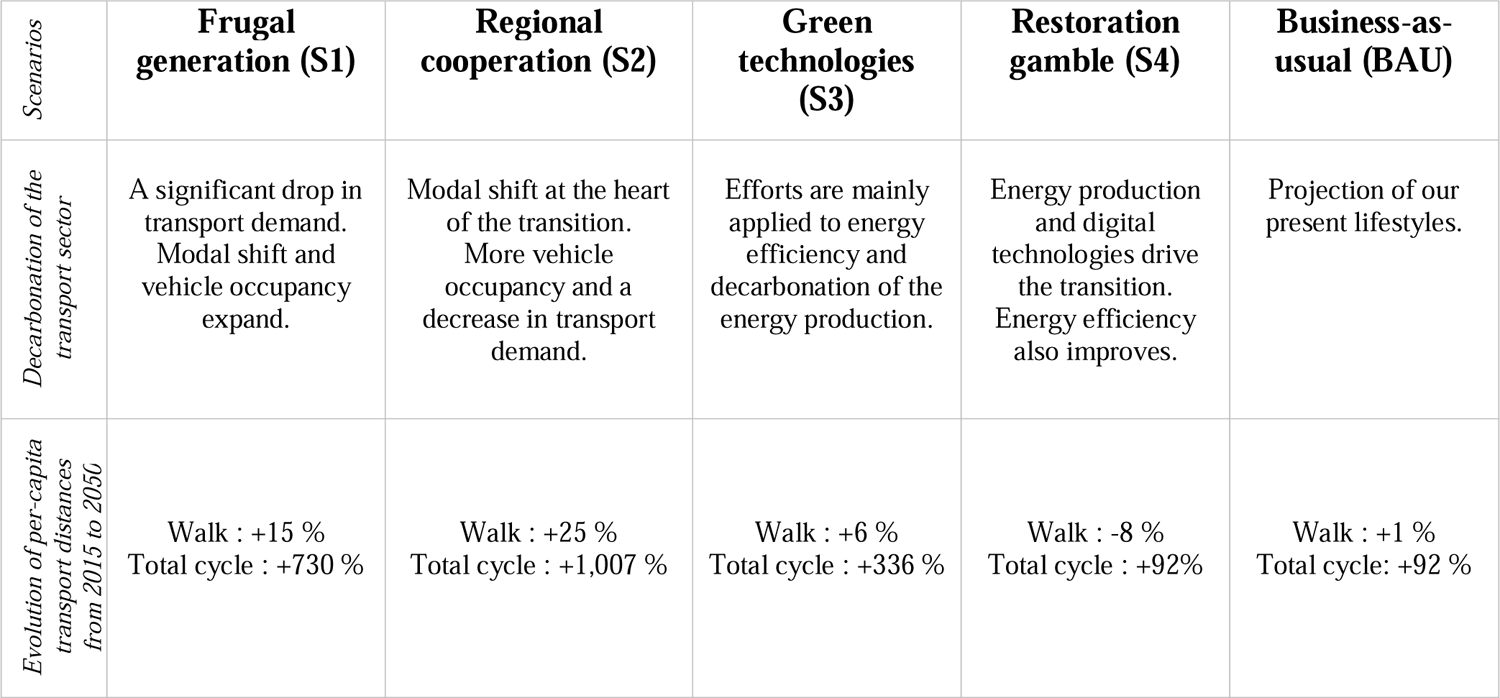
Transport sector and active mobility in ADEME’s prospective scenarios.

### Age-specific allocation of active transport demands

For each scenario and mode of transportation, the passenger-kilometres were distributed among age groups according to the mode-specific age distributions observed in the 2019 French mobility survey (Ministry of Ecological Transition, n.d.).

In addition, we allocated the total number of kilometres cycled between E-bikes and mechanical bicycles differently based on age groups. This allocation was done in order to achieve two objectives: i) replicate the observed 6.7-year age difference between E-bikers and cyclists as reported in a recent multi-country study (Castro et al., 2019), and ii) reflect the changing relative contribution of E-bikes to the total cycling distance over time, as assumed in each scenario. Further details regarding these calculations are provided in the Supplementary Materials.

### Demographic projections

Population projections, including mortality and life expectancy, for different years and age groups, were accessed from publicly available data provided by the French national institute of statistics and economic studies (INSEE, n.d.). These same data were also used by ADEME in the design of all their transition scenarios.

### Health impact assessment

We relied on international guidelines to model and report the health effects associated with active transportation of the *Transitions2050* scenarios for metropolitan France over the 2021-2050 period (Hess et al., 2020). Our health modelling method relies on principles similar to the HEAT tool developed by WHO (Götschi et al., 2020), but differs in two major aspects. First, we only explicitly considered physical activity for the pathway linking active transport and health. Second, we accounted for the age-distribution of current levels of active transport in France and for the age-difference between e-bike and classical bike users.

For each age group, yearly distances travelled were converted into exposure times for walking, cycling or E-cycling using the following average speeds, respectively: 4.8, 14.9 and 18.1 km.h^-1^ (Egiguren et al., 2021; Götschi et al., 2020). First, we compared the resulting total duration of active travel to international recommendations for weekly moderate physical activity, averaged over the entire adult population.

The health impacts (number of deaths averted) generated by physical activity associated with active transportation were calculated, using a linear dose-response function between duration spent in active transportation and the reduction in all-cause mortality, weighted by the specific mortality rate and size of the population:

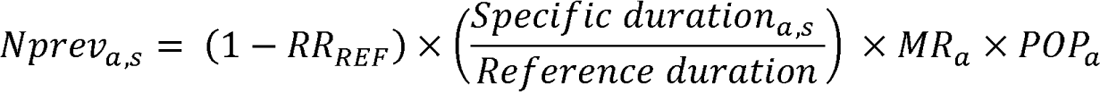

With:

*Nprev_a,s_*: Number of death prevented

*RR*_REF_: Relative risk of all-cause mortality associated with the reference duration of active travel (Kelly et al., 2014).

*Specific duration*_a,s_: Projected in the scenarios and distributed across age groups

*Reference duration*: 168 minutes for walking and 100 minutes for cycling as estimated in a meta-analysis of 18 cohorts (Kelly et al., 2014). This correspond to an energy expenditure of 11.25 MET.hour.week^-1^.

*MR_a_*: Specific mortality rate of an age categorie *a*

*POP_a_*: Population of the age category *a*

We used a relative risk (RR_REF_) of 0.89 for all-cause mortality when engaging weekly in 168 minutes of walking, with maximum risk reduction of 30%, and a relative risk of 0.90 for 100 minutes of cycling, with a maximum risk reduction of 45%. Health impacts are therefore capped above 460 minutes of walking and 447 minutes of cycling (Kelly et al., 2014). We constrained the benefits associated with physical activity to the population aged between 20 and 89 years. This restriction was made based on the fact that the relative risk estimates for cycling and walking were extrapolated from studies encompassing individuals within the age range of 20 to 93 years (Kelly et al., 2014) and the data used for age distribution of cycling distances were collected from individuals aged between 15 and 89 years (Ministry of Ecological Transition, n.d.). The assumed ratio of energy expenditure between moderate assistance E-bikes and bikes was 0.89, as estimated by a recent meta-analysis (McVicar et al., 2022). The impacts were evaluated for each age group, taking into account the respective population size and mortality rate. The total count of deaths prevented by each scenario was obtained, together with the number of years of life lost (YLL) prevented, by subtracting from the estimated number of deaths prevented in this scenario the estimated number of deaths prevented in the BAU scenario. Finally, based on the estimate of the number of deaths prevented, life tables were generated in order to compute the life expectancy in each scenario.

### Monetizing the health impacts

We further estimated the intangible costs associated with the health impact of each scenario, using the value of a statistical life year (VSLY) that is recommended in France for the socioeconomic assessment of public investments (Emile, 2013). In France, the VSLY is projected to be €151,000 in 2025, and this figure is anticipated to increase to €202,000 by 2050.

The VSLY was multiplied by the number of YLL prevented to calculate the monetary impacts of mortality risk reduction. All monetarized values were expressed in €_2022_.

All the estimates were presented alongside uncertainty intervals (UI), which were determined using the lower and upper bounds of the 95% confidence interval (CI) of the RR used.

### Sensitivity analyses

We performed five distinct sensitivity analyses. First, we explored a more favourable RR associated with cycling (0.81 instead of 0.90 per 100 minutes of cycling), as estimated by a recent meta-analysis (Zhao et al., 2021). Second, we explored alternative walking and cycling speeds, respectively 3.6 and 13 km.h^-1^ (The French Agency for Ecological Transition, n.d.). Third, we investigated an alternative ratio of 0.78 between E-bike and cycling energy expenditures (Berntsen et al., 2017). Fourth, for scenarios S1 and S2, we assumed that the high increase of cycling overall would affect the demography of cyclist toward a flatter age distribution, as those observed in countries with a high cycling culture, such as Denmark. We therefore evolved gradually between 2021 to 2035 the distribution of age-specific contribution of kilometres cycled from the levels observed in France (Ministry of Ecological Transition, n.d.) to those reported in Denmark (Center for Transport Analytics, Transport, n.d.) (see supplementary materials). Lastly, we constrained the benefits of active transportation to the age range of 20 to 75 years, as opposed to the wider scope of 20 to 89 years in the main analysis.

## Results

### Projection of active transport

Time changes in transport demand from 2021 to 2050 are presented in supplementary materials (Figure S1) for each mode of active transport and scenario. Figure 2 compares the estimated average weekly durations of active transportation obtained under each scenarios by 2035 and 2050 with both the 2015 French situation and the minimal recommendations set by the World Health Organization (WHO), that is, 150 minutes of moderate-intensity aerobic physical activity per week. In 2015, the active transportation levels in the French population allowed for a weekly average of 80 minutes of moderate-intensity aerobic physical activity (The French Agency for Ecological Transition, n.d.). In the S2 scenario, in 2035, the average levels of physical activity through active transportation alone are expected to attain the minimal WHO recommendations. The S1 scenario nearly achieves these levels by 2035 and surpasses them by 2050. In contrast, the S3 scenario falls short of reaching the minimal recommended levels of physical activity, both in the medium term (2035) and the long term (2050). Both the BAU scenario and the S4 scenario project minimal increases in active transportation compared to 2015, and fail to reach WHO’s minimal recommendations, even by 2050. These results are detailed by age groups in supplementary materials (Figure S2).

**Figure 2.**
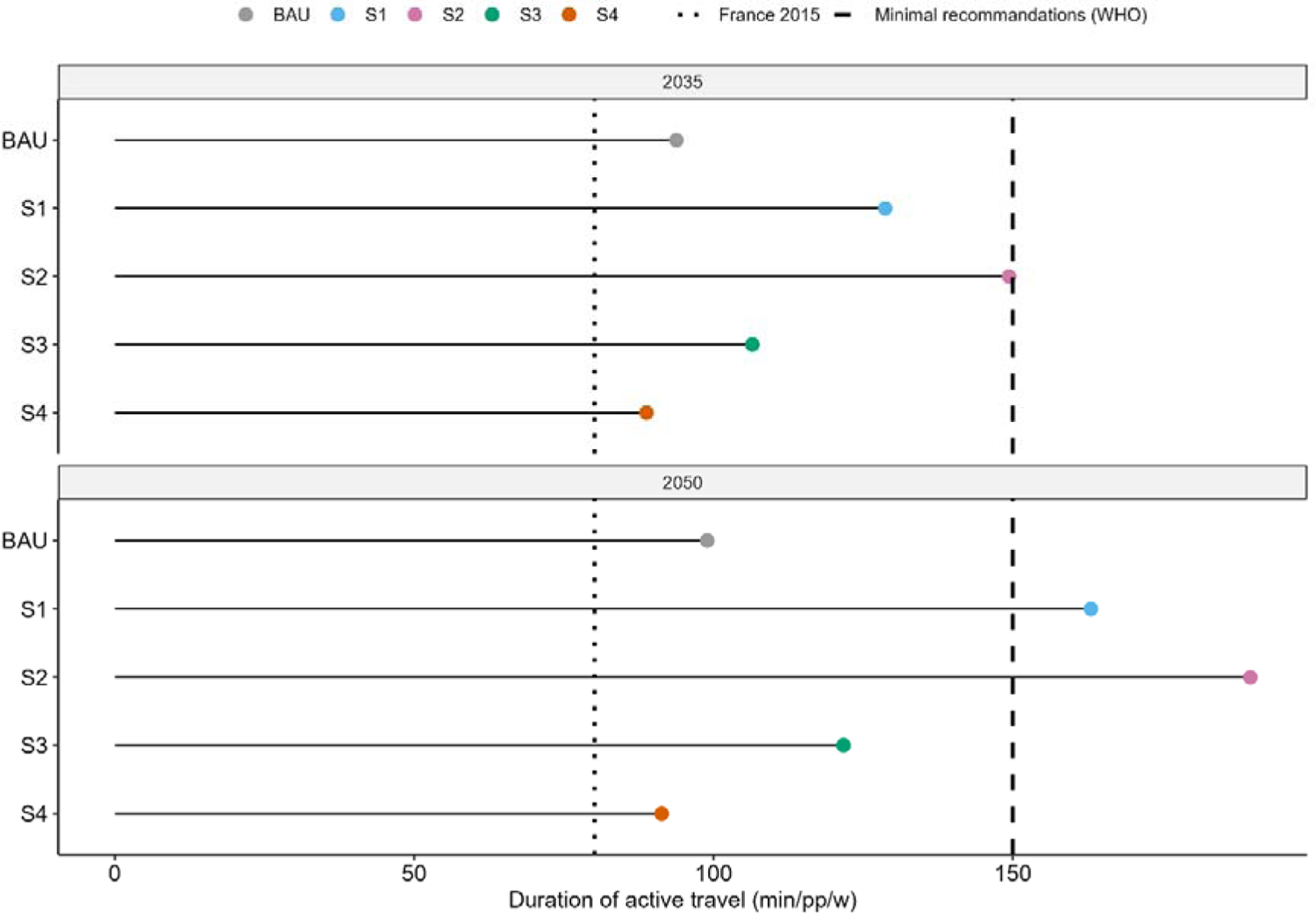
Duration (averaged for the 20-89 years old) of physical activity generated by active transportation in 2035 and 2050 compared to the French levels in 2015 (all estimated by ADEME Transition scenarios) and the WHO guidelines for moderate physical activity [26].

### Quantitative health impact assessment

The time-varying health effects expected in each scenario are presented in Figure 3 as numbers of prevented deaths and YLL compared to the BAU. The disparities in health impacts among scenarios become more pronounced as time progresses. From 2030 to 2040, the benefits of S1 and S2 scenarios experience a large increase, while the benefits of S3 scenario show a more linear progression. On the other hand, S4 scenario has negative impacts that intensify over time and remain noticeable for all age groups above 50 years old. Deaths prevented by scenarios S1-S3 are highest in the 80-89 age group, and YLL prevented increase for younger ages, plateau for the 40-59 and the 60-79 years old age groups, and are limited amongst 80-89 years old.

**Figure 3.**
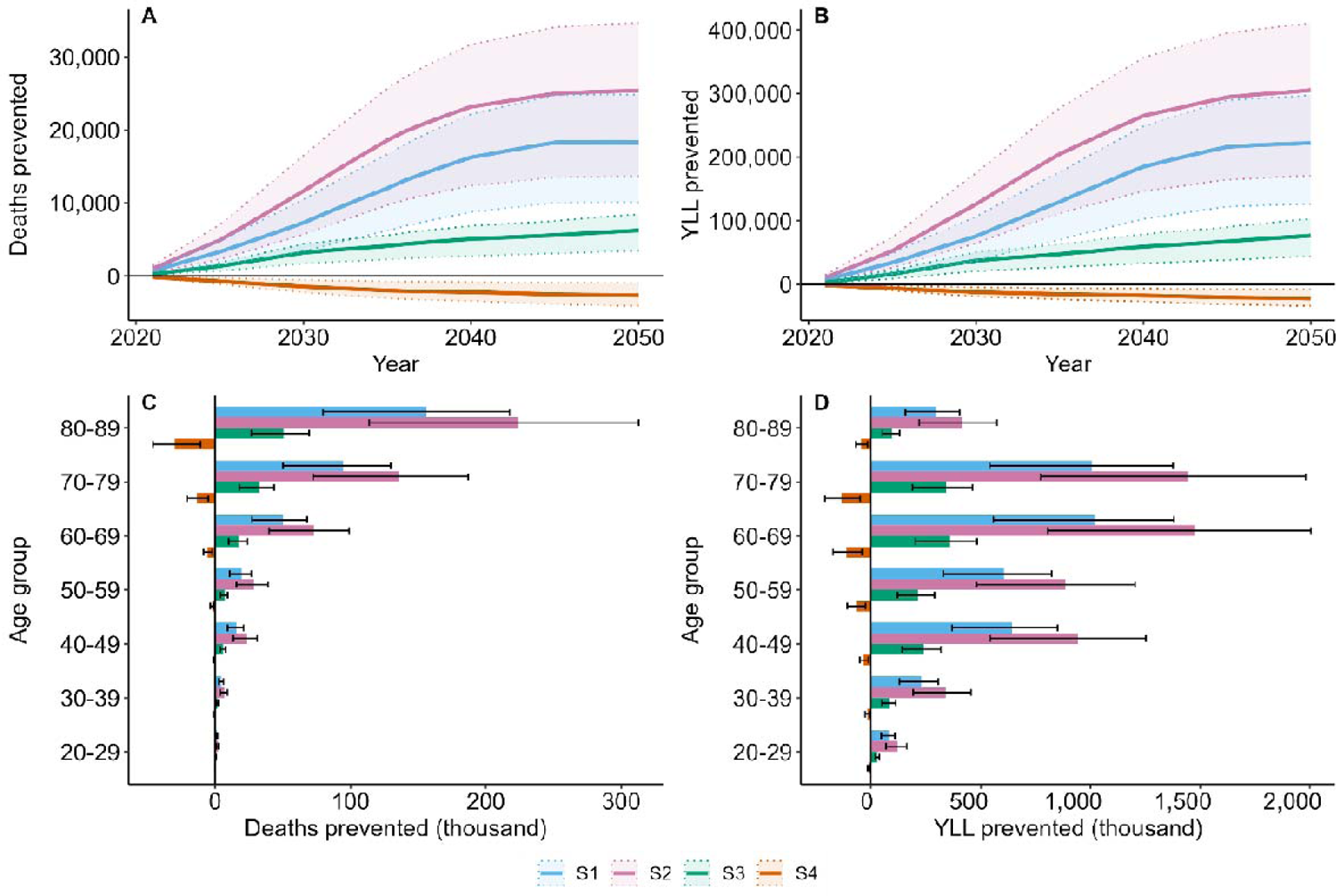
Deaths prevented (a) and (c) and YLL prevented (b) and (d) by year ((a) and (b)) and age group ((c) and (d)), cumulative for the 2021-2050 period) for each transition scenario compared to the BAU.

For all age groups, the benefits of S2 surpass all other scenarios while the benefits of S1 outweigh those of S3. The YLL metric makes these benefits apparent for younger age groups (from 20 years old on). The health impacts of S4 remain small, though negative.

Figure 4 distinguishes the health benefits of the different scenarios by type of active transportation. Starting from 2030 onwards, scenarios S1 and S2 demonstrate a rapid increase in the health benefits associated with E-cycling, while its progression in S3 is more linear. The health benefits in the first scenario mainly come from cycling, with more or less equal contributions of classical and E-cycling throughout the period from 2021 to 2050. In scenarios S2 and S3, E-cycling emerges as the mode of active transportation contributing to the majority of health benefits, while walking and cycling provide similar benefits each year. On the other hand, scenario S4, which is comparable to the BAU scenario in terms of cycling mobility, exhibits lower levels of walking. Consequently, the health detriments observed in scenario S4 can be entirely attributed to the decrease in pedestrian mobility.

**Figure 4.**
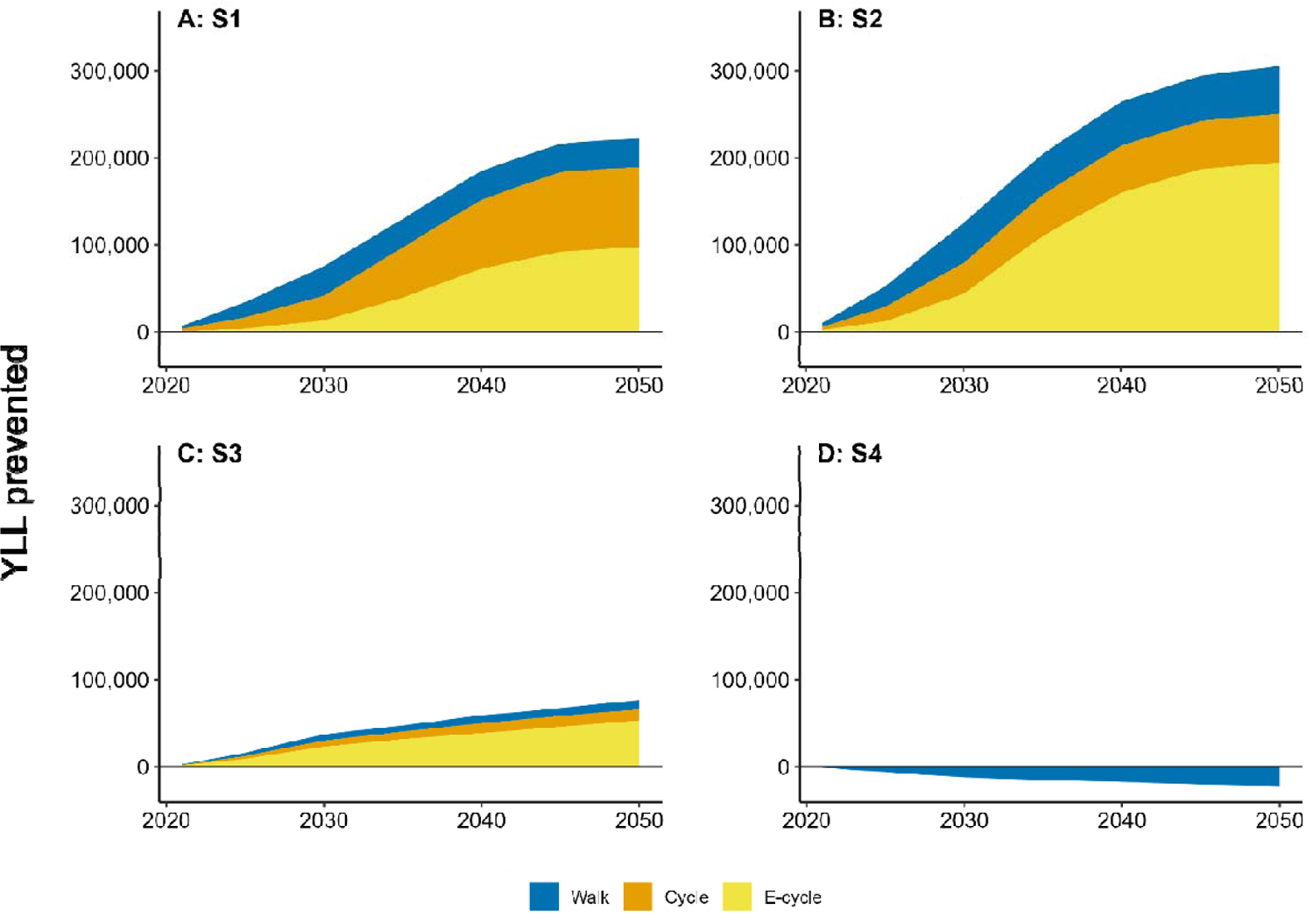
Years of Life Lost (YLL) prevented as a function of time and type of active transportation, under (a) scenario S1, (b) scenario S2, (c) scenario S3, and (d) scenario S4.

Figure 5 presents the annual health and economic benefits of each scenario at medium (2035) and long (2050) term. Here again, scenario S2 shows a larger impact over all other scenarios, S1 exhibits a superiority over scenario S3, and scenario S4 has detrimental health impacts as compared to the BAU. The number of deaths prevented in 2050 by active transportation reaches 25,402 [13,687-34,661] with the trajectories projected for scenario S2, 18,345 [10,083-24,833] with scenario S1 and 6,250 [3,443-8,469] with scenario S3. Scenario S4 would induce an additional 2,658 [962-4,106] deaths.

**Figure 5.**
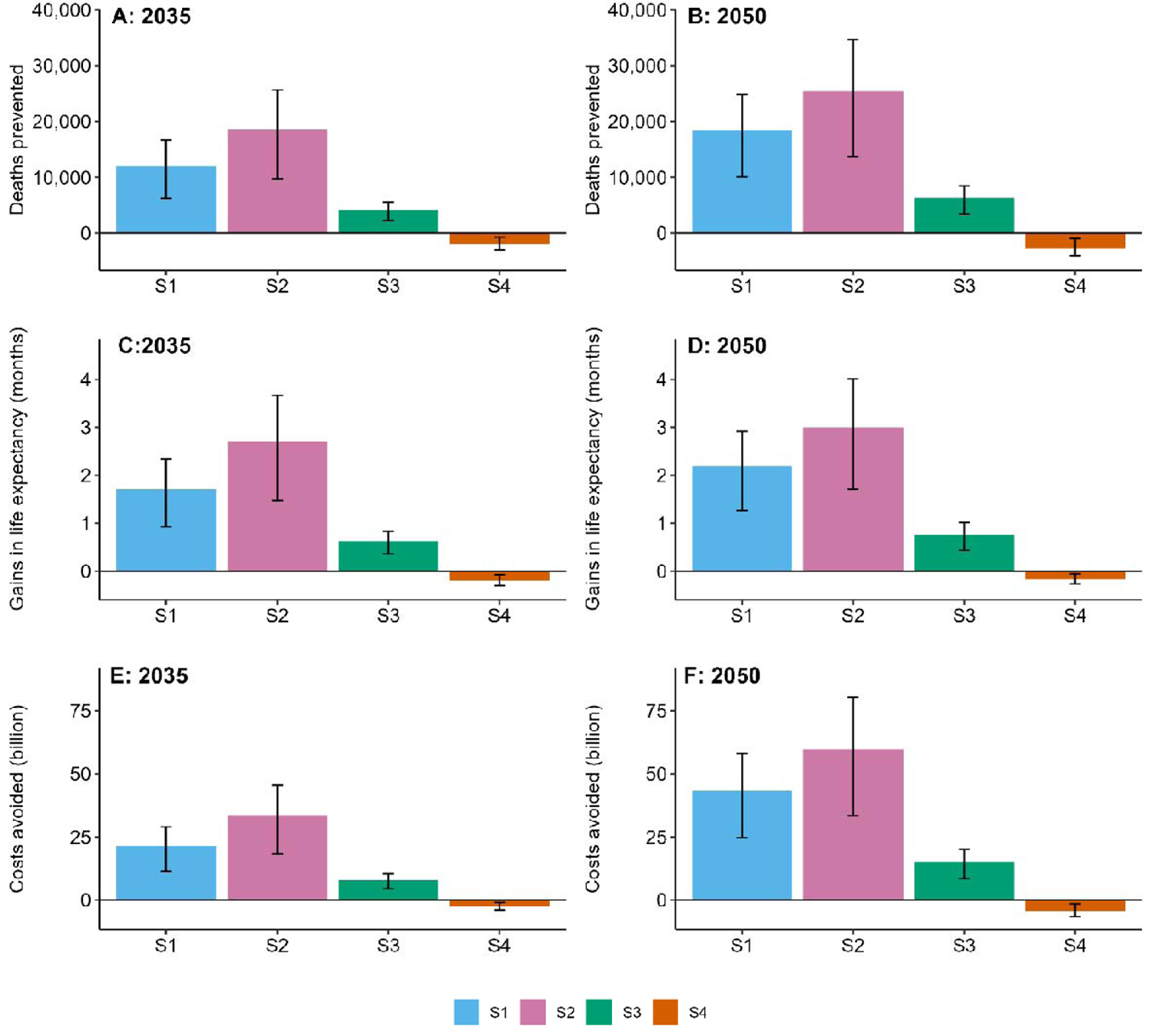
Deaths prevented ((a) and (b)), gain in life expectancy ((c) and (d)) and cost avoided ((e) and (f)) by 2035 ((a), (c), (e)) and 2050 ((b), (d), (f)).

Life expectancy of the French population in 2050 would improve by 3.0 [1.7-4.0] months by achieving modal shift as described in scenario S2, 2.2 [1.3-2.9] months with scenario S1, 0.8 [0.4-1.0] months with scenario S3 and would decrease by 0.2 [0.1-0.3] months with scenario S4.

For 2035, the annual corresponding monetarized health benefits (based on the YLL) range from €33.7 [IC: 18.3-45.5] billion with scenario S2 to €7.8 [IC: 4.5-10.4] billion with scenario S3. Scenario S4 corresponds to a net detrimental monetarized health impact of €2.5 [1.0-3.9] billion.

### Sensitivity Analyses

Table 2 summarizes the results of our sensitivity analyses. Overall, the parameter choice which affects the most our estimates is the RR value for cycling. Assuming a more favourable RR leads to notable increases in the YLL estimated, around +50 to +70%. Modifying the speeds of walking and cycling has less but substantial effects of about +20% in the YLL prevented. By lowering the speeds, the duration of exposure to physical activity is extended, leading to higher health impacts. The third sensitivity analysis, focusing on the ratio of energy expenditure provided by an E-bike compared to a mechanical one slightly reduces the health impact, by less than 10% of YLL prevented. In scenarios characterized by high levels of cycling (S1 and S2), using the gradual shape of the age distribution of cycling kilometres of France in 2021 to Denmark in 2035 (and on to 2050) leads to a minor decrease in the number of prevented YLL. Considering Denmark’s relative age contribution for cycling kilometres leads to less deaths prevented in older age groups (Supplementary materials, figure S5), and so has limited impacts on YLL prevented. Restricting the health effects to individuals below the age of 75 reduces the YLL prevented ascribed to active transportation by about 20%.

**Table 2.**
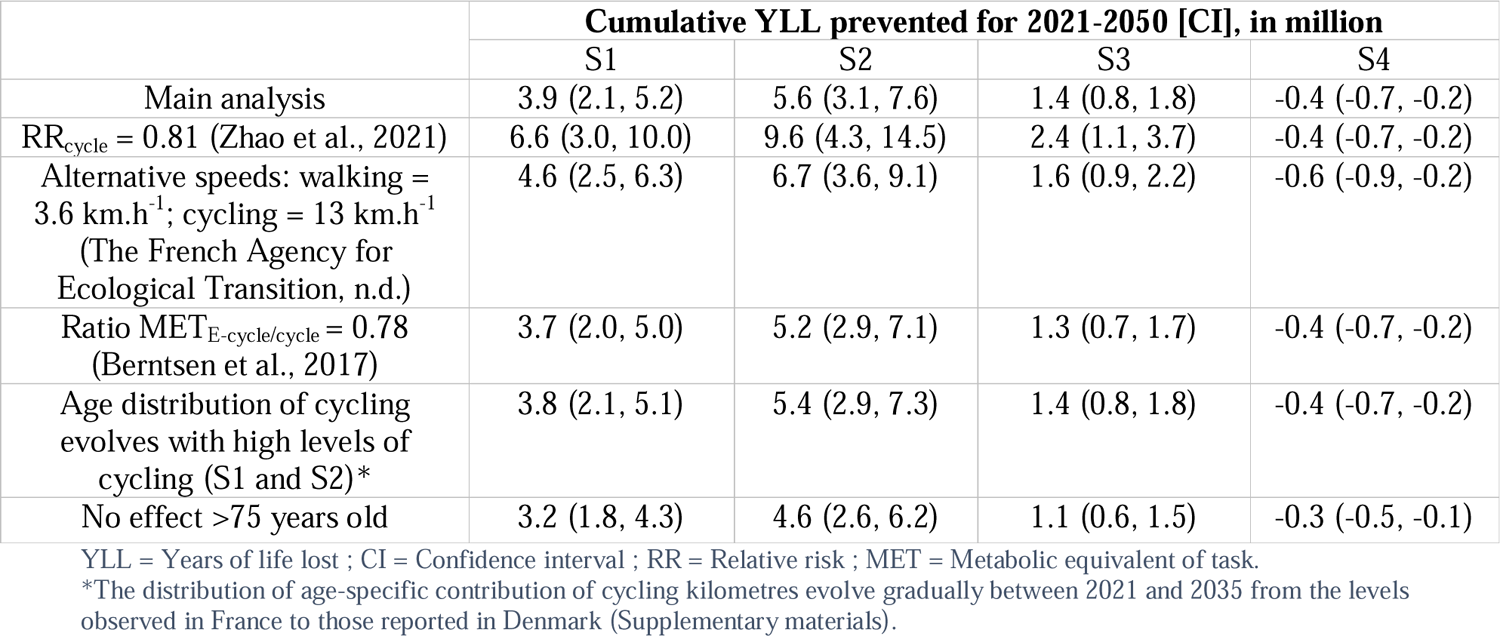
Health impacts of active transportation for ADEME’s scenarios. Main and sensitivity analysis.

|  | Cumulative YLL prevented for 2021-2050 [CI], in million |  |  |  |
| --- | --- | --- | --- | --- |
|  | S1 | S2 | S3 | S4 |
| Main analysis | 3.9 (2.1, 5.2) | 5.6 (3.1, 7.6) | 1.4 (0.8, 1.8) | -0.4 (-0.7, -0.2) |
| RR<sub>cycle</sub> = 0.81 (Zhao et al., 2021) | 6.6 (3.0, 10.0) | 9.6 (4.3, 14.5) | 2.4 (1.1, 3.7) | -0.4 (-0.7, -0.2) |
| Alternative speeds: walking = 3.6 km.h<sup>-1</sup>; cycling = 13 km.h<sup>-1</sup><br>(The French Agency for Ecological Transition, n.d.) | 4.6 (2.5, 6.3) | 6.7 (3.6, 9.1) | 1.6 (0.9, 2.2) | -0.6 (-0.9, -0.2) |
| Ratio MET<sub>E-cycle/cycle</sub> = 0.78<br>(Berntsen et al., 2017) | 3.7 (2.0, 5.0) | 5.2 (2.9, 7.1) | 1.3 (0.7, 1.7) | -0.4 (-0.7, -0.2) |
| Age distribution of cycling evolves with high levels of cycling (S1 and S2)* | 3.8 (2.1, 5.1) | 5.4 (2.9, 7.3) | 1.4 (0.8, 1.8) | -0.4 (-0.7, -0.2) |
| No effect >75 years old | 3.2 (1.8, 4.3) | 4.6 (2.6, 6.2) | 1.1 (0.6, 1.5) | -0.3 (-0.5, -0.1) |

YLL = Years of life lost ; CI = Confidence interval ; RR = Relative risk ; MET = Metabolic equivalent of task.
\*The distribution of age-specific contribution of cycling kilometres evolve gradually between 2021 and 2035 from the levels observed in France to those reported in Denmark (Supplementary materials).

For each analysis and any choices of the parameters, the ranking of scenarios according to their health impacts remained unchanged, with S2 as the most beneficial scenario, followed by S1 and then S3, while S4 yielded negative health impacts.

## Discussion

### Main results

In this study, we highlight the widely varying potential health benefits of transport transition pathways towards carbon neutrality, through their impact on active transport. Future pathways promoting modal shift to active modes of transportation facilitate the attainment of the minimal recommendation on physical activity set by the WHO solely through transport activities. Conversely, policies wagering on future technological developments to attain carbon neutrality are likely to intensify the lack of physical activity and its harmful health impacts in the general population. According to the extent of modal shift, and compared to the BAU, health impacts at the 2035 horizon may range from 19,000 deaths prevented to 2,000 additional deaths, representing respectively €34 billion annual savings and €3 billion additional mortality costs. Qualitative results and the ranking of *Transitions2050* scenarios remained consistent through several sensitivity analyses.

### Literature comparison

For all scenarios, the majority of health benefits may be explained by an achievable increase in cycling mobility, particularly through the use of E-bikes. The most optimistic scenario reaches 20 kilometres cycled and 7 kilometres walked per person weekly, representing roughly 20 minutes per day of active transportation. Such increase in active transportation seem considerable as compared to the levels reported before 2020. However, they remain lower to those reported in other European countries: In Netherlands, active transportation represent between 24 and 28 minutes daily (Fishman et al., 2015) and more than 50% of trips <2 kilometres and 30% of trips between 2 to 5 kilometres are undertaken using bicycles (Goel et al., 2021). This highlights the margin of progression for active transportation in France, where only 5% of the active population uses a bike for trips covering less than 5 kilometres, while 60% opt for a car (Chantal Brutel, Jeanne Pages, 2021). The anticipated levels of active transportation in S1 and S2 scenarios therefore appear to be quite achievable.

The method we used for quantifying health and economic impacts follows the recommended steps and parameters outlined by the WHO and environmental economists. These approaches are widely described and employed in various scientific studies (Emile, 2013; Götschi et al., 2020; Smith et al., 2021). A health impact assessment study on the transport sector was previously conducted in France using a similar method (Barban et al., 2022). The scenario explored in this study projected active transportation levels similar to those observed in the S1 scenario, and the estimated health impacts were similar to those we obtain. Studies comparing multiple scenarios consistently demonstrate significant differences in the health impacts of active transportation: Climate policy strategies that prioritize health-promoting pathways and are built upon the behaviours of the population yield more significant health benefits (Hamilton et al., 2021; Milner et al., 2023).

Preventing, as we predict in scenario S2, 19,000 deaths annually by 2035 solely through deliberate decisions in the development of the transportation sector holds significant importance for public health prevention as it corresponds to 2.8% of all-cause mortality projected for this year (INSEE, n.d.). Indeed, in comparison, in France, it is estimated that exposure to PM2.5 is responsible for 40,000 deaths annually (Santé publique France, 2021) while heat-attributable deaths reached up to 7,000 in 2022 (Santé publique France, 2023). In the transport sector, all road-related fatalities in France range between 3,000 and 4,000 deaths annually (excepted in 2020 and 2021) (ONISR (Observatoire national interministériel de la sécurité routière), 2023). From a benefit-cost perspective, based on a UK case-study, the economic gain of prevented mortality generated by an additional 267 regular cyclists per kilometres of a new separated cycleway breaks even on the costs of construction (Candio and Frew, 2023).

The adoption of electric modes of transportation is widely recognized as a strategy for mitigating emissions within the transportation sector. However, it is important to acknowledge that electric vehicles, along with the high battery metals demand (European Federation for Transport and Environment, 2023), can also contribute to pollutant emissions through the dispersion of road dust particles and the degradation of tires and brakes wear (Bourliva et al., 2017). In the transport sector, modal shift and decarbonation of the energy supply can yield larger health co-benefits (Milner et al., 2023). The promotion of E-bikes is a cost-effective approach to reduce transport associated carbon emissions while improving health and mobility (Jenkins et al., 2022). The allocation of more areas to bicycle infrastructure can also optimize the utilization of public space (Gössling et al., 2016).

Other outcomes have to be taken into account when outlining health effects of modal shift such as air pollution, road injuries or noise. As the health risk estimate relies on a prospective cohort, these factors are implicitly incorporated into the relative risks. Regardless of the specific assumptions, the health benefits of active transportation consistently outweigh the potential adverse effects stemming from exposure to air pollution and traffic-related incidents (Mueller et al., 2018). Among various modes of transportation, car users exhibit highest levels of exposure to air pollutants such as PM2.5, black carbon, and CO (de Nazelle et al., 2017). The association with road injuries depends greatly on the assumptions made on the fatality and injury rates of cyclists (Quam et al., 2017) as well as the quality of transport infrastructures (WHO, 2022). Furthermore, each incremental shift from cars to bicycles corresponds to a noteworthy decrease in pollutant emissions (Bernard et al., 2021). For instance, it has been estimated that an individual who replaces one car trip per day with a bicycle trip over 200 days can reduce their CO_2_ emissions by 0.5 tons per year (Brand et al., 2021). Noise pollution would also benefit from modal shift, which would yield positive impacts on health-related quality of life, mediated by annoyance and sleep disturbance (Héritier et al., 2014). It is also expected that a general increase of physical activity reduces morbidity related to cardiovascular risks, cancer, osteoporosis, depression and low back pain (Miller et al., 2016). Further impacts related to quality of life can also benefit from active transportation such as work performance and productivity (Ding et al., 2016).

### Limitation

Using the transport demand as a proxy for physical activity allows for a quantification of health impacts associated with specific active modes of transport but does not encompass several variations in the population. For example, given the expectation that scenarios S3 and S4 would limit active transportation primarily to those who are already physically active, while scenarios S1 and S2 would promote active transportation for a broader population, including individuals who are sedentary, a more targeted distribution of transport demand could be achieved. This distribution would involve a heterogeneous allocation of distances within age groups. By implementing such an allocation strategy, the differences in health impacts between high cycling (S1, S2) and low cycling (S3, S4) scenarios could be amplified. Another limitation linked to this proxy is that the physical activities performed out of transport context are not taken into account, despite their anticipated distinct evolution across scenarios. However, several studies suggest that active transportation adds to rather than replaces other physical activities (Laeremans et al., 2017).

Using a relative risk from a meta-analysis provides a robust association between physical activity and mortality reduction but does not allow to specify distinct relative benefits based on age, gender, or socio-occupational category. A recent meta-analysis showed no significant differences in the association of cycling with all-cause mortality for gender groups and among individuals aged above or below 50 years (Zhao et al., 2021). Additional cohort studies are needed to assess the potential heterogeneity of the association between more specific age groups and socio-occupational categories.

Our analysis does not explicitly take into account the potential adverse effects of cycling and walking, such as crashes and exposure to air pollution. However, since our assessment relies on a dose-response function linking physical activity and all-cause mortality, these harmful effects are implicitly considered. In addition, it is very likely that our analysis involves an overestimation of the adverse effects of traffic-related pollution and crashes, especially in scenarios identified as the most favourable (S1 and S2). Indeed, in S1 and S2, the kilometres travelled by cars are assumed to decrease dramatically, and substantially more than in TEND, S3, and S4 (Table S1). In Europe, differences in overall crash rates between cities are driven largely by crashes that involved motor-vehicles (Branion-Calles et al., 2020). This reduction of car traffic in S1 and S2 would also lead to a decrease in the exposure of pedestrians and cyclists to traffic-related air pollution. At last, these scenarios involve a substantial increase in bike volumes (Figure S1) while evidence suggests that the individual risk of fatal crashes during walking and cycling decreases as the volumes of active transportation increase (Jacobsen et al., 2015; WHO, 2022).

Our results are calculated from a projected scenario, the BAU. This inherent uncertainty may introduce variability in the absolute results of each scenario, yet it does not alter the relative hierarchy of their respective impacts.

All in all, it is important to acknowledge that a health impact assessment study only provides theoretical estimates of health and economic impacts, and that its quantitative results should not be over-interpreted. However, the magnitude of effects and the comparison of scenarios allow for a better interpretation of the co-benefits and potential implications of specific public policies.

## Conclusion

Different carbon neutral trajectories imply distinct health benefits, this holds particularly true within the transport sector. The implementation of public policies promoting modal shift effectively improves population health while reducing the use of fossil fuels and pollutant emissions. In France, directing transportation towards active modes would yield significant health co-benefits. Transition trajectories primarily relying on technological interventions offer minimal benefits and may even exacerbate the lack of physical activity in the population. In a context of reaching towards carbon neutrality, these results may help guide decision-makers in the choice of optimal transition strategies.

## Data availability statements

All analyses were conducted using the R software version 4.2.3; all data and codes are publicly available at: https://github.com/LeoMoutet/HIA-transport.

## Acknowledgments

We are thankful to Stephane Barbusse (ADEME - The French Agency for Ecological Transition) for useful elements and discussions regarding the ADEME *Transitions2050* scenarios.

## Authors contributions

PQ and KJ conceived the design of this study. LM produced output, figures and tables. AB contributed to data acquisition. LM, LT and KJ interpreted the results and wrote the original draft of the article. AB and PQ contributed to critical feedbacks on the methodology and interpretation of the results.

