## Supplementary materials for "Different pathways toward net-zero emissions imply diverging health impacts: a health impact assessment study for France"

### **Distribution of kilometres travelled for electric bicycles**

ADEME specified the number of kilometres travelled according to the type of bicycle (conventional vs electric). For each year and scenario, we can therefore calculate the proportion $\Pi$ of kilometres travelled on electric bicycles. It was 5% in 2015 and 23% in 2020 for all scenarios. By 2050, this proportion reaches 60% for S1, 80% for S2 and S3, and 75% for S4 and TEND. These proportions are therefore targeted for 2050, let us note them "$obj\_\Pi$."

The proportion of users is not equivalent between conventional and electric bicycles according to age. An average difference of 6.7 years was estimated between users of conventional vs electric bicycles (Castro et al., 2019), we note this difference as $\delta$. Therefore, it is necessary to estimate the vector $\pi_{a}$ representing the proportion of kilometres travelled on electric bicycles for each age group. This vector must satisfy two conditions: firstly, the sum of its values applied to each age group must lead to the proportion of electric bicycle users in the general population,$\Pi$.

Moreover: 0 < $\pi_{a}$ < 1; if $\pi_{a}$< 0, it is assigned the value 0, and if $\pi_{a}$> 1, it is assigned the value 0.99.

That is:

$$\frac{\sum_{a} {pop}_{a}\times{km\_pp}_{a}\times\pi_{a}}{KM}=\Pi$$

Secondly, the age difference between users of conventional bicycles and e-bikes, weighted by the distances travelled by each age group, must be equal to $\delta$, which is 6.7.

Thus:

$\frac{\sum_{a} a\times{km\_pp}_{a}\times\pi_{a}}{\sum_{a} {km\_pp}_{a}\times\pi_{a}}$ **-** $\frac{\sum_{a} a\times{km\_pp}_{a}\times{(1-\pi}_{a})}{\sum_{a} {km\_pp}_{a}\times(1-\pi_{a})}\boldsymbol{=}\delta$

We want $\Pi$ and $\delta$ as:

$$5\times\left( \Pi-obj\_\Pi\right)+\left( \delta-obj\_\delta\right)\to0$$

With $obj\_\delta$ = 6.7 and for S1, S2, S3, S4, TEND, respectively, obj_$\Pi$ = 0.6, 0.8, 0.8, 0.75, 0.75

We assume that $\pi_{a}$ is defined linearly according to age, and we use the R optimization function to obtain the parameters x and y:

$$\pi_{a}=x\times a+y$$

### **Distribution of kilometres travelled per year by age (Sensitivity analyse 4)**

For a sensitivity analysis, in order to distribute the distances travelled in the population for high cycling scenarios (S1 and S2), transport demands were allocated for each five-year age group according to the relative contribution of each age group observed in France for 2021 (Ministry of Ecological Transition, n.d.), and linearly evolve to the Danish levels (Center for Transport Analytics, Transport, n.d.) in 2035 and on to 2050.

The number of kilometres travelled per person per year depends on the contribution of the age group to the total kilometres travelled according to the proportions observed in France or Denmark. In practice, the first age group of interest (15-19 years) served as the reference, and the ratio of the number of kilometres travelled per person to the reference group was computed for each age group.

Let us note: ${km\_pp}_{a}^{Den}$ the number of kilometres travelled by age group $a$.

The first age group of interest, 15-19 year olds, is use as the reference, and then for each age group, the ratio $\rho$ of the number of kilometres travelled per person compared to the reference group is calculated:

$$\forall a \in\left( 2,n \right), \rho_{a}^{Den}=\frac{{km\_pp}_{a}^{Den}}{{km\_pp}_{1}^{Den}}$$

With $\forall a \in\left( 2,n \right), {km\_pp}_{a}^{Fr}= \rho_{a}\times$ ${km\_pp}_{1}^{Fr}$

The ratios for each age must correspond to the ratios of those same ages in France. This in order to obtain a French distribution that matches the Danish one independently of the kilometres travelled per year and the age structure of the population:

$$\forall a \in\left( 1,n \right), \rho_{a}^{Den}= \rho_{a}^{Fr}\leftrightarrow\frac{{km\_pp}_{a}^{Den}}{{km\_pp}_{1}^{Den}}=\frac{{km\_pp}_{a}^{Fr}}{{km\_pp}_{1}^{Fr}}$$

For each year, we have to calculate the value${km\_pp}_{a}^{Fr}$, which, for each age group, must match the distribution of French relative transport demands with that observed in Denmark. This is independent of the age structure, noted${pop}_{a}^{Fr}$, and representing the number of person in each age group$a$.

The transport demands projected by ADEME are expressed in passenger- kilometres (${KM}^{Fr}$), that is, the total number of kilometres travelled by the population (${pop}_{a}^{Fr}$) in a year, as follows:

$${KM}^{Fr}=\sum_{a} {pop}_{a}^{Fr}\times{km_{pp}}_{a}^{Fr}$$

That is:	${KM}^{Fr}={km\_pp}_{1}^{Fr}\times\left( {pop}_{1}^{Fr}+ \sum_{a=2}^{n} \rho_{a}\times{pop}_{a}^{Fr} \right)$

Therefore:	${km\_pp}_{1}^{Fr}= \frac{{KM}^{Fr}}{{pop}_{1}^{Fr}+ \sum_{a=2}^{n} \rho_{a}\times{pop}_{a}^{Fr}}$ $\leftrightarrow\boldsymbol{km\_pp}_{\boldsymbol{k}}^{\boldsymbol{Fr}}\boldsymbol{=}\frac{\boldsymbol{\rho}_{\boldsymbol{k}}\boldsymbol{\times}\boldsymbol{KM}^{\boldsymbol{Fr}}}{\boldsymbol{pop}_{\boldsymbol{1}}^{\boldsymbol{Fr}}\boldsymbol{+}\sum_{\boldsymbol{a=2}}^{\boldsymbol{n}} \boldsymbol{\rho}_{\boldsymbol{a}}\boldsymbol{\times}\boldsymbol{pop}_{\boldsymbol{a}}^{\boldsymbol{Fr}}}$

### **Evolution of age distributions (Sensitivity analyse 4)**

For each scenario to be evaluated (S1 and S2), there are two elements that differ from the baseline scenario used for comparison (the demographic structure or the applied mortality rates remain the same between the BAU and evaluated scenarios):

- The absolute level of overall physical activity $B_{t}$, which varies over time t.
- The relative distribution of this overall level among age classes, which may or may not vary over time: $p\left( a,t \right)$, and which adheres to $\sum_{a} p(a,t)=1$ regardless of t.

The evaluation time horizon is defined as 2035.

If we assume a variation in both the levels and the age distribution of physical activity, then a target distribution $p_{c}$ needs to be defined (e.g., that of the Denmark) to be achieved for a targeted year $t_{c}$, 2035:

$$p\left( a,t_{c} \right)= p_{c}(a)$$

The initial age distribution (in 2021) is denoted as $p\left( a,t_{0} \right)$.

For all $t$ such that $t_{0}<t<t_{c}$, we can assume that the intermediate age distribution is defined as follows:

$$p\left( a,t \right)=p\left( a,t_{0} \right)+\left( p_{c}\left( a \right)-p\left( a,t_{0} \right) \right)\times\frac{(t-t_{0})}{{(t}_{c}-t_{0})}$$

This relative distribution, varying over time, is used at each intermediate year to distribute the variable overall absolute level of physical activity $B_{t}$, which is also variable over time.

### **Relative contribution of age groups (Sensitivity analyse 4)**

The allocation of kilometres travelled according to age groups is a crucial step in estimating the health impacts. For Denmark’s pyramid, the distribution of kilometres cycled across different age groups closely follows the population size, whereas the observed distribution in France exhibits a bimodal pattern, with a higher allocation of kilometres to older individuals. The distribution of kilometres walked is slightly different, French population from 55 years old are attributed an increased proportion of kilometres while Denmark’s distribution shows a relative steady distribution across all ages.

In Denmark, no available data was available for individuals aged from 85 to 89 years old. To address this data gap, we applied a proportional adjustment, similar to the relative levels observed from the 75-79 age group as compare to the 80-84 age group.

| *Scenarios* | **Frugal generation (S1)** | **Regional cooperation (S2)** | **Green technologies (S3)** | **Restoration gamble (S4)** | **Business-as-usual (BAU)** |
| --- | --- | --- | --- | --- | --- |
| *Decarbonation of the transport sector* | A significant drop in transport demand. Modal shift and vehicle occupancy expand. | Modal shift at the heart of the transition.  More vehicle occupancy and a decrease in transport demand. | Efforts are mainly applied to energy efficiency and decarbonation of the energy production. | Energy production and digital technologies drive the transition. Energy efficiency also improves. | Projection of our present lifestyles. |
| *Modal share of active transport in 2050* | 51.1% | 47.7% | 31.6% | 20.0% | 22.4% |
| *Share of e-bike in the km travelled by bicycle (%)* | 2030: 0.4  2050: 0.6 | 2030: 0.55  2050: 0.8 | 2030: 0.6  2050: 0.8 | 2030: 0.45  2050: 0.75 | 2030: 0.45  2050: 0.75 |
| *Evolution of per-capita transport distances from 2015 to 2050* | Walk : +15 %  Total cycle : +730 %  Train: +20 %  Car: -49%  Bus: +3% | Walk : +25 %  Total cycle : +1,007 %  Train: +45 %  Car: -40%  Bus: +22% | Walk : +6 %  Total cycle : +336 %  Train: +26 %  Car: -8%  Bus: +27% | Walk : -8 %  Total cycle : +92%  Train: 18 %  Car: -3%  Bus: +13% | Walk : +1 %  Total cycle: +92 %  Train: 15 %  Car: +11%  Bus: +10% |
| *Minutes spent per day per capita for each type of transport in 2050* | Walk: 16.4  Total cycle: 11.7  Train: 3.3  Car: 16.7  Bus and coach: 2.6 | Walk: 17.7  Total cycle: 13.6  Train: 3.8  Car: 18.7  Bus and coach: 3.1 | Walk: 15.1  Total cycle: 6.2  Train: 3.2  Car: 26.3  Bus and coach: 3.3 | Walk: 13.1  Total cycle: 2.5  Train: 2.9  Car: 27.6  Bus and coach: 2.9 | Walk: 14.4  Total cycle: 2.9  Train: 2.8  Car: 31.6  Bus and coach: 2.8 |

**Table S1: Transport sector and general mobility in ADEME’s prospective scenarios**

**Table S2: Deaths prevented in the main and sensitivity analyses, cumulated for the 2021-2050 period and annually for 2035 and 2050:**

|  | S1 | S2 | S3 | S4 |
| --- | --- | --- | --- | --- |
|  | **Cumulative deaths prevented for 2021-2050 (CI), in thousand** | | | |
| Main analysis | 342 (180, 471) | 494 (260, 681) | 116 (63, 157) | -52 (-81, -19) |
| RR_cycle_ = 0.81 (Zhao et al., 2021) | 558 (246, 844) | 802 (353, 1214) | 197 (88, 298) | -52 (-81, -19) |
| Alternative speeds: walking = 3.6 km.h^-1^; cycling = 13 km.h^-1^ (The French Agency for Ecological Transition, n.d.) | 412 (214, 571) | 597 (309, 827) | 138 (75, 188) | -70 (-108, -25) |
| Ratio MET_E-cycle/cycle_ = 0.78 (Berntsen et al., 2017) | 328 (172, 442) | 464 (241, 641) | 107 (58, 146) | -52 (-81, -19) |
| Age distribution of cycling evolves with high levels of cycling (S1 and S2)* | 238 (118, 335) | 354 (176, 499) | 116 (63, 157) | -52 (-81, -19) |
| No effect >75 years old | 136 (75, 184) | 198 (109, 268) | 49 (27, 65) | -14 (-22, -5) |
|  | **Deaths prevented in 2035 (CI), in thousand** | | | |
| Main analysis | 12 (6, 17) | 19 (10, 26) | 4 (2, 6) | -2 (-3, -1) |
| RR_cycle_ = 0.81 (Zhao et al., 2021) | 19 (8, 29) | 30 (13, 45) | 7 (3, 11) | -2 (-3, -1) |
| Alternative speeds: walking = 3.6 km.h^-1^; cycling = 13 km.h^-1^ (The French Agency for Ecological Transition, n.d.) | 15 (7, 20) | 22 (12, 31) | 5 (3, 7) | -3 (-4, -1) |
| Ratio MET_E-cycle/cycle_ = 0.78 (Berntsen et al., 2017) | 12 (6, 16) | 18 (9, 24) | 4 (2, 5) | -2 (-3, -1) |
| Age distribution of cycling evolves with high levels of cycling (S1 and S2)* | 8 (4, 12) | 13 (6, 19) | 4 (2, 6) | -2 (-3, -1) |
| No effect >75 years old | 5 (3, 7) | 8 (4, 10) | 2 (1, 2) | -1 (-1, 0) |
|  | **Deaths prevented in 2050 (CI), in thousand** | | | |
| Main analysis | 18 (10, 25) | 25 (14, 35) | 6 (3, 8) | -3 (-4, -1) |
| RR_cycle_ = 0.81 (Zhao et al., 2021) | 31 (14, 47) | 43 (19, 64) | 11 (5, 16) | -3 (-4, -1) |
| Alternative speeds: walking = 3.6 km.h^-1^; cycling = 13 km.h^-1^ (The French Agency for Ecological Transition, n.d.) | 22 (12, 30) | 30 (16, 42) | 7 (4, 10) | -4 (-5, -1) |
| Ratio MET_E-cycle/cycle_ = 0.78 (Berntsen et al., 2017) | 17 (10, 24) | 24 (13, 32) | 6 (3, 8) | -3 (-4, -1) |
| Age distribution of cycling evolves with high levels of cycling (S1 and S2)* | 12 (6, 16) | 17 (9, 24) | 6 (3, 8) | -3 (-4, -1) |
| No effect >75 years old | 7 (4, 9) | 10 (5, 13) | 2 (1, 3) | -1 (-1, 0) |

* The distribution of age-specific contribution of cycling kilometres evolve gradually between 2021 and 2035 from the levels observed in France to those reported in Denmark.

**Table S3: Years of life lost prevented (YLL) in the main and sensitivity analyses, cumulated for the 2021-2050 period and annually for 2035 and 2050:**

|  | S1 | S2 | S3 | S4 |
| --- | --- | --- | --- | --- |
|  | **Cumulative YLL prevented for 2021-2050 (CI), in million** | | | |
| Main analysis | 3.9 (2.1, 5.2) | 5.6 (3.1, 7.6) | 1.4 (0.8, 1.8) | -0.4 (-0.7, -0.2) |
| RR_cycle_ = 0.81 (Zhao et al., 2021) | 6.6 (3.0, 10.0) | 9.6 (4.3, 14.5) | 2.4 (1.1, 3.7) | -0.4 (-0.7, -0.2) |
| Alternative speeds: walking = 3.6 km.h^-1^; cycling = 13 km.h^-1^ (The French Agency for Ecological Transition, n.d.) | 4.6 (2.5, 6.3) | 6.7 (3.6, 9.1) | 1.6 (0.9, 2.2) | -0.6 (-0.9, -0.2) |
| Ratio MET_E-cycle/cycle_ = 0.78 (Berntsen et al., 2017) | 3.8 (2.1, 5.1) | 5.4 (2.9, 7.3) | 1.4 (0.8, 1.8) | -0.4 (-0.7, -0.2) |
| Age distribution of cycling evolves with high levels of cycling (S1 and S2)* | 3.8 (2.1, 5.1) | 5.4 (3.0, 7.3) | 1.4 (0.8, 1.9) | -0.4 (-0.7, -0.2) |
| No effect >75 years old | 3.2 (1.8, 4.3) | 4.6 (2.6, 6.2) | 1.1 (0.6, 1.5) | -0.3 (-0.5, -0.1) |
|  | **YLL prevented in 2035 (CI), in thousand** | | | |
| Main analysis | 130 (70, 177) | 205 (111, 277) | 48 (27, 63) | -15 (-24, -6) |
| RR_cycle_ = 0.81 (Zhao et al., 2021) | 218 (97, 330) | 349 (156, 526) | 85 (38, 127) | -15 (-24, -6) |
| Alternative speeds: walking = 3.6 km.h^-1^; cycling = 13 km.h^-1^ (The French Agency for Ecological Transition, n.d.) | 155 (83, 212) | 244 (132, 332) | 56 (31, 75) | -20 (-32, -7) |
| Ratio MET_E-cycle/cycle_ = 0.78 (Berntsen et al., 2017) | 125 (67, 171) | 191 (103, 260) | 44 (25, 58) | -15 (-24, -6) |
| Age distribution of cycling evolves with high levels of cycling (S1 and S2)* | 127 (68, 172) | 197 (106, 267) | 48 (27, 63) | -15 (-24, -6) |
| No effect >75 years old | 108 (59, 147) | 172 (95, 232) | 40 (23, 53) | -12 (-18, -4) |
|  | **YLL prevented in 2050 (CI), in thousand** | | | |
| Main analysis | 222 (126, 297) | 305 (171, 411) | 77 (44, 103) | -22 (-34, -8) |
| RR_cycle_ = 0.81 (Zhao et al., 2021) | 395 (179, 596) | 534 (240, 806) | 137 (61, 207) | -22 (-34, -8) |
| Alternative speeds: walking = 3.6 km.h^-1^; cycling = 13 km.h^-1^ (The French Agency for Ecological Transition, n.d.) | 261 (147, 350) | 361 (200, 488) | 90 (51, 122) | -29 ( -45, -11) |
| Ratio MET_E-cycle/cycle_ = 0.78 (Berntsen et al., 2017) | 210 (119, 281) | 281 (157, 380) | 70 (40, 95) | -22 (-34, -8) |
| Age distribution of cycling evolves with high levels of cycling (S1 and S2)* | 209 (118, 280) | 284 (158, 383) | 77 (44, 103) | -22 (-34, -8) |
| No effect >75 years old | 174 (100, 232) | 239 (136, 320) | 61 (35, 81) | -15 (-23, -5) |

* The distribution of age-specific contribution of cycling kilometres evolve gradually between 2021 and 2035 from the levels observed in France to those reported in Denmark.

**Table S4: Gain in life expectancy in the main and sensitivity analyses, annually for 2035 and 2050:**

|  | S1 | S2 | S3 | S4 |
| --- | --- | --- | --- | --- |
|  | **Gain in life expectancy in 2035 (CI), in months** | | | |
| Main analysis | 1.7 (0.9, 2.3) | 2.7 (1.5, 3.7) | 0.6 (0.4, 0.8) | -0.2 (-0.3, -0.1) |
| RR_cycle_ = 0.81 (Zhao et al., 2021) | 2.9 (1.3, 4.4) | 4.6 (2.1, 7.0) | 1.1 (0.5, 1.7) | -0.2 (-0.3, -0.1) |
| Alternative speeds: walking = 3.6 km.h^-1^; cycling = 13 km.h^-1^ (The French Agency for Ecological Transition, n.d.) | 2.0 (1.1, 2.8) | 3.2 (1.7, 4.4) | 0.7 (0.4, 1.0) | -0.3 (-0.4, -0.1) |
| Ratio MET_E-cycle/cycle_ = 0.78 (Berntsen et al., 2017) | 1.6 (0.9, 2.3) | 2.5 (1.4, 3.4) | 0.6 (0.3, 0.8) | -0.2 (-0.3, -0.1) |
| Age distribution of cycling evolves with high levels of cycling (S1 and S2)* | 1.7 (0.9, 2.3) | 2.6 (1.4, 3.5) | 0.6 (0.4, 0.8) | -0.2 (-0.3, -0.1) |
| No effect >75 years old | 1.7 (0.9, 2.3) | 2.7 (1.5, 3.7) | 0.6 (0.4, 0.8) | -0.2 (-0.3, -0.1) |
|  | **Gain in life expectancy in 2050 (CI), in months** | | | |
| Main analysis | 2.2 (1.3, 2.9) | 3.0 (1.7, 4.0) | 0.8 (0.4, 1.0) | -0.2 (-0.3, -0.1) |
| RR_cycle_ = 0.81 (Zhao et al., 2021) | 4.0 (1.8, 6.0) | 5.4 (2.4, 8.1) | 1.4 (0.6, 2.1) | -0.2 (-0.3, -0.1) |
| Alternative speeds: walking = 3.6 km.h^-1^; cycling = 13 km.h^-1^ (The French Agency for Ecological Transition, n.d.) | 2.6 (1.5, 3.4) | 3.5 (2.0, 4.7) | 0.9 (0.5, 1.2) | -0.2 (-0.3, -0.1) |
| Ratio MET_E-cycle/cycle_ = 0.78 (Berntsen et al., 2017) | 2.1 (1.2, 2.7) | 2.8 (1.6, 3.7) | 0.7 (0.4, 0.9) | -0.2 (-0.3, -0.1) |
| Age distribution of cycling evolves with high levels of cycling (S1 and S2)* | 2.3 (1.3, 3.1) | 3.1 (1.8, 4.1) | 0.8 (0.4, 1.0) | -0.2 (-0.4, -0.1) |
| No effect >75 years old | 2.2 (1.3, 2.9) | 3.0 (1.7, 4.0) | 0.8 (0.4, 1.0) | -0.2 (-0.3, -0.1) |

* The distribution of age-specific contribution of cycling kilometres evolve gradually between 2021 and 2035 from the levels observed in France to those reported in Denmark.

**Table S5: Costs avoided in the main and sensitivity analyses, cumulated for the 2021-2050 period and annually for 2035 and 2050:**

|  | S1 | S2 | S3 | S4 |
| --- | --- | --- | --- | --- |
|  | **Cumulative costs avoided for 2021-2050 (CI), in billion** | | | |
| Main analysis | 679 (374, 919) | 983 (539, 1330) | 238 (135, 319) | -73 (-113, -26) |
| RR_cycle_ = 0.81 (Zhao et al., 2021) | 1168 (523, 1772) | 1683 (752, 2560) | 422 (190, 642) | -73 (-113, -26,) |
| Alternative speeds: walking = 3.6 km.h^-1^; cycling = 13 km.h^-1^ (The French Agency for Ecological Transition, n.d.) | 807 (439, 1096) | 1169 (634, 1590) | 281 (157, 377) | -97 (-35, -150) |
| Ratio MET_E-cycle/cycle_ = 0.78 (Berntsen et al., 2017) | 649 (356, 878) | 914 (498, 1240) | 219 (123, 294) | -73 (-113, -26) |
| Age distribution of cycling evolves with high levels of cycling (S1 and S2)* | 657 (361, 888) | 938 (512, 1271) | 238 (135, 319) | -73 (-113, -26) |
| No effect >75 years old | 554 (308, 746) | 803 (445, 1083) | 197 (113, 263) | -53 (-82, -19) |
|  | **Costs avoided in 2035 (CI), in billion** | | | |
| Main analysis | 21 (12, 29) | 34 (18, 46) | 8 (4, 10) | -3 (-4, -1) |
| RR_cycle_ = 0.81 (Zhao et al., 2021) | 36 (16, 54) | 57 (26, 87) | 14 (6, 21) | -3 (-4, -1) |
| Alternative speeds: walking = 3.6 km.h^-1^; cycling = 13 km.h^-1^ (The French Agency for Ecological Transition, n.d.) | 25 (14, 35) | 40 (22, 55) | 9 (5, 12) | -3 (-5, -1) |
| Ratio MET_E-cycle/cycle_ = 0.78 (Berntsen et al., 2017) | 21 (11, 28) | 31 (17, 43) | 7 (4, 10) | -3 (-4, -1) |
| Age distribution of cycling evolves with high levels of cycling (S1 and S2)* | 21 (11, 28) | 32 (18, 44) | 8 (4, 10) | -3 (-4, -1) |
| No effect >75 years old | 18 (10, 24) | 28 (16, 38) | 7 (4, 9) | -2 (-3, -1) |
|  | **Costs avoided in 2050 (CI), in billion** | | | |
| Main analysis | 43 (25, 58) | 60 (33, 80) | 15 (9, 20) | -4 (-7, -2) |
| RR_cycle_ = 0.81 (Zhao et al., 2021) | 77 (35, 116) | 104 (47, 158) | 27 (12, 40) | -4 (-7, -2) |
| Alternative speeds: walking = 3.6 km.h^-1^; cycling = 13 km.h^-1^ (The French Agency for Ecological Transition, n.d.) | 51 (29, 68) | 70 (39, 95) | 18 (10, 24) | -6 (-9, -2) |
| Ratio MET_E-cycle/cycle_ = 0.78 (Berntsen et al., 2017) | 41 (23, 55) | 55 (31, 74) | 14 (8, 18) | -4 (-7, -2) |
| Age distribution of cycling evolves with high levels of cycling (S1 and S2)* | 41 (23, 55) | 55 (31, 75) | 15 (9, 20) | -4 (-7, -2) |
| No effect >75 years old | 34 (20, 45) | 47 (27, 63) | 12 (7, 16) | -3 (-4, -1) |

* The distribution of age-specific contribution of cycling kilometres evolve gradually between 2021 and 2035 from the levels observed in France to those reported in Denmark.

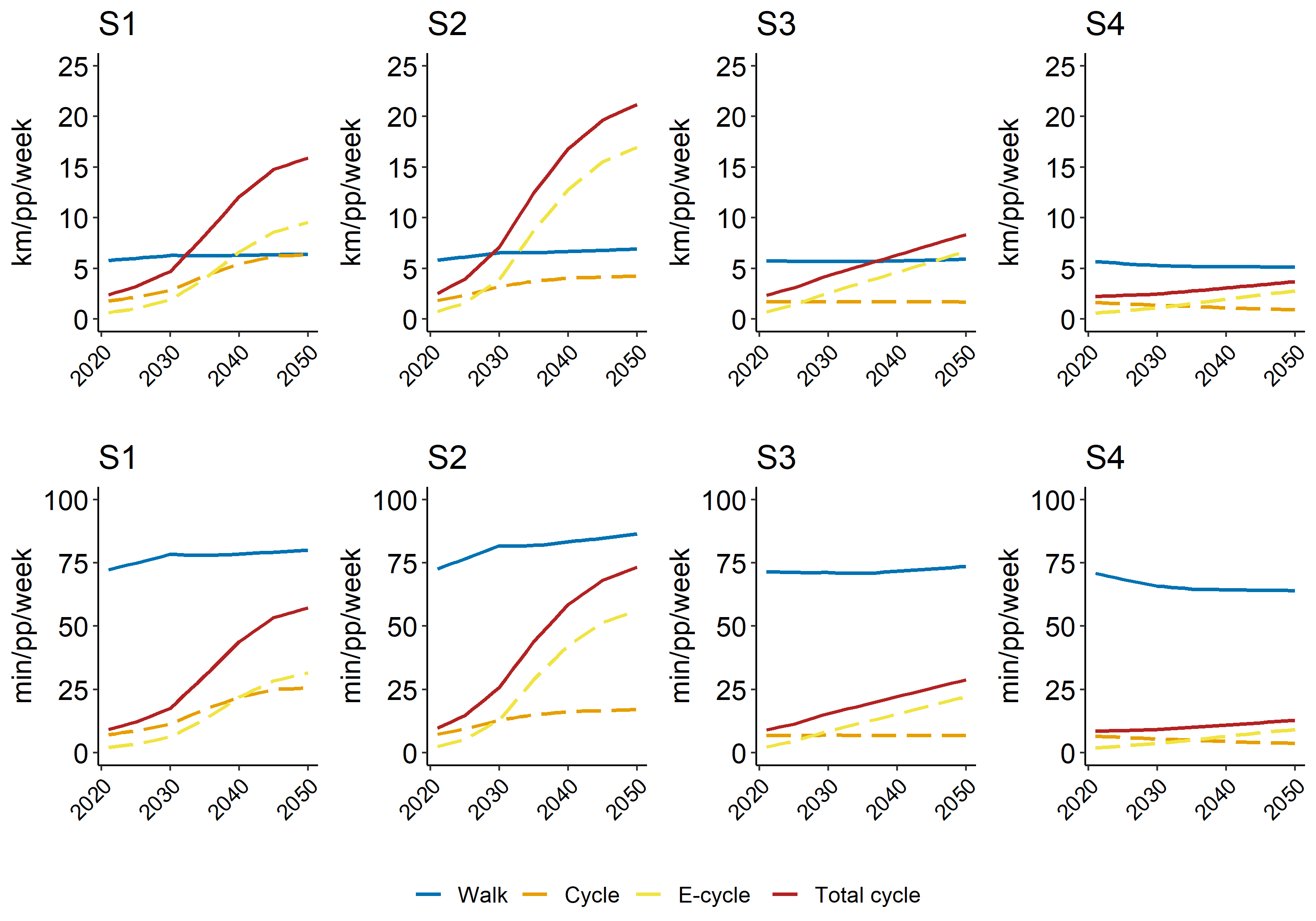

**Figure S1: Evolution of transport demand from 2021 to 2050 for each mode of active transportation.**

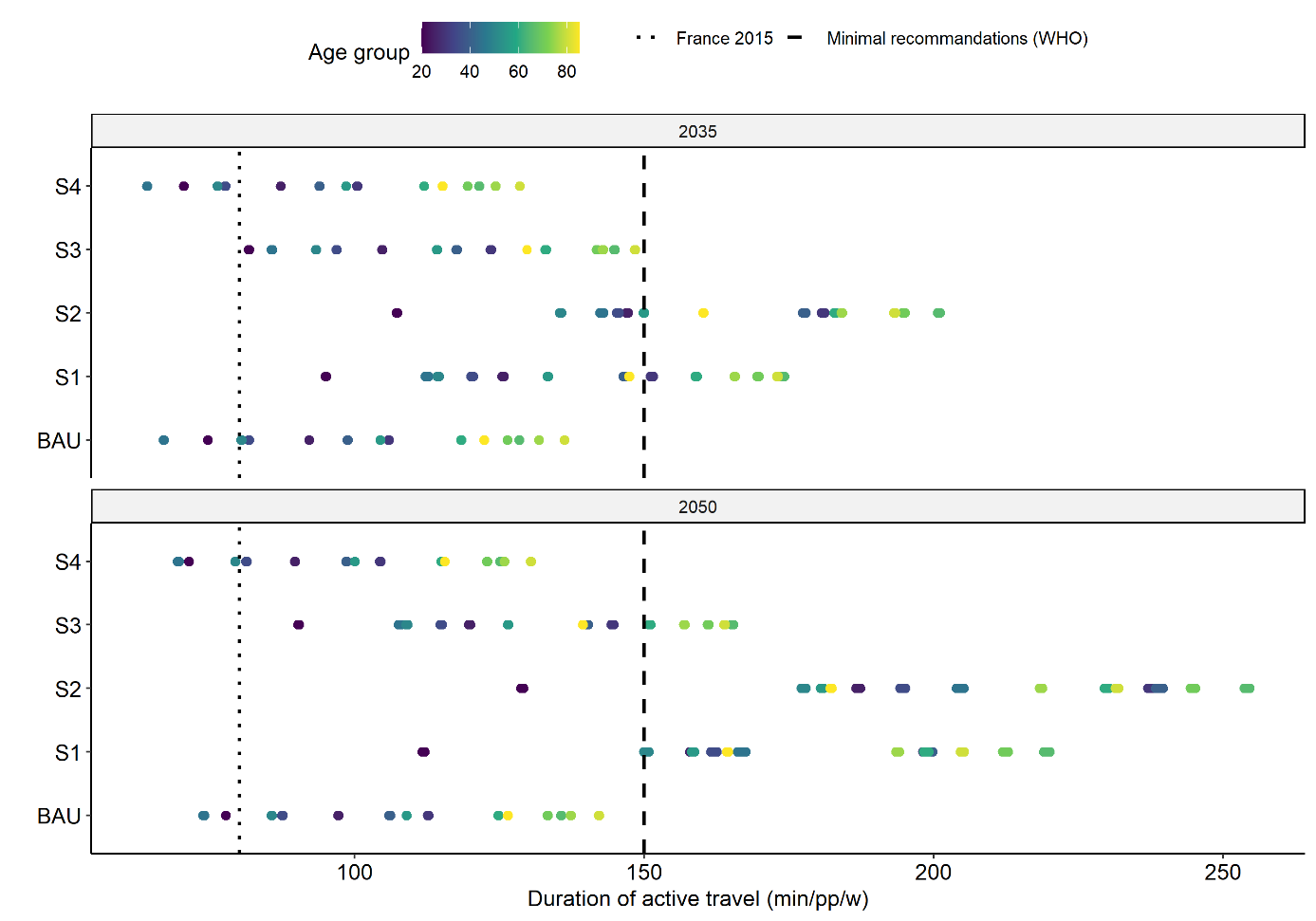

**Figure S2: Duration of physical activity generated by active transportation in 2035 and 2050 compared to the French levels in 2015 (all estimated by ADEME Transition scenarios) and the WHO guidelines for moderate physical activity by age groups.**

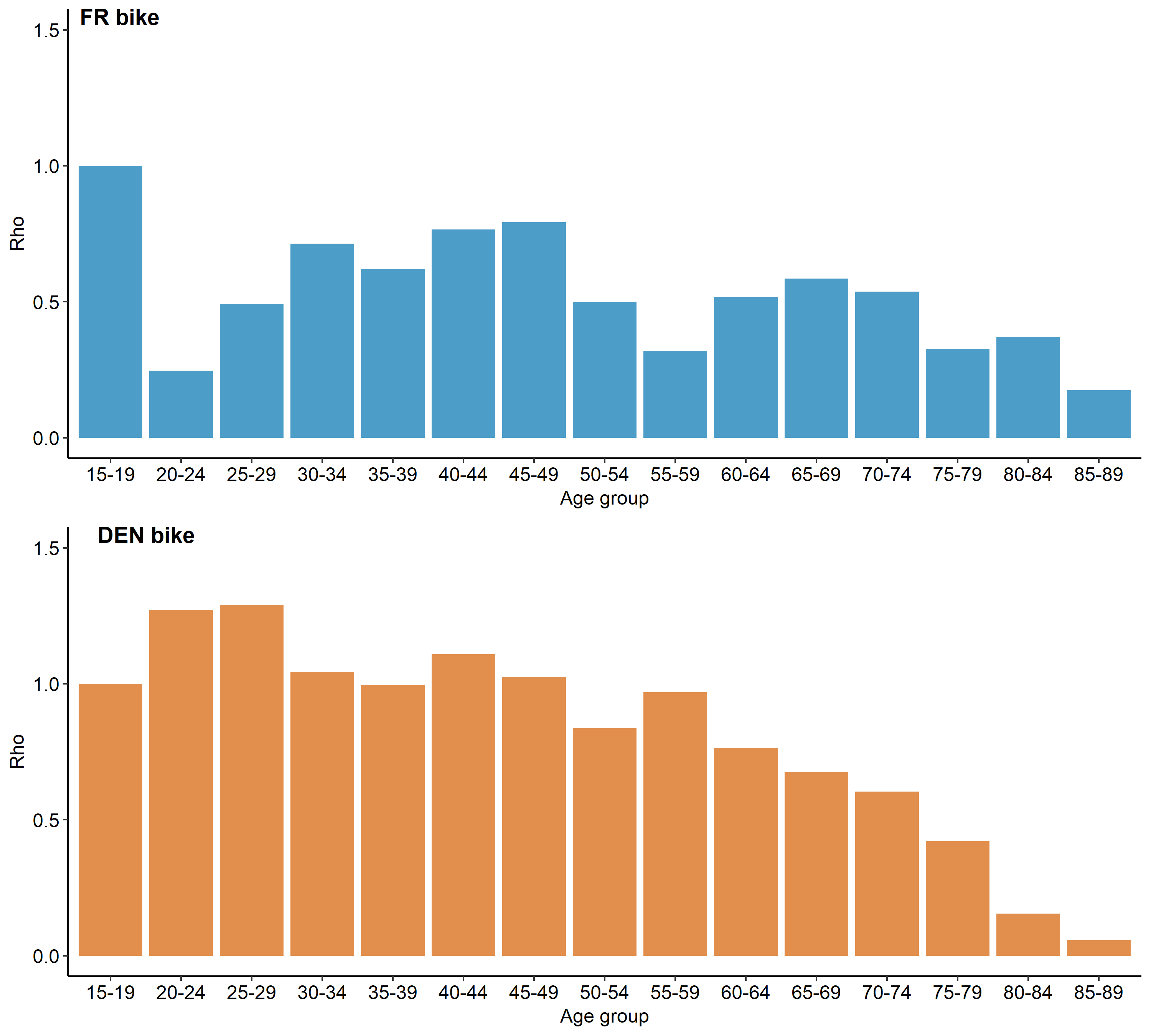

**Figure S3: Age distribution of cycling distances across age groups (from 15-19 to 85-89 years) in France (blue) and Denmark (orange):**

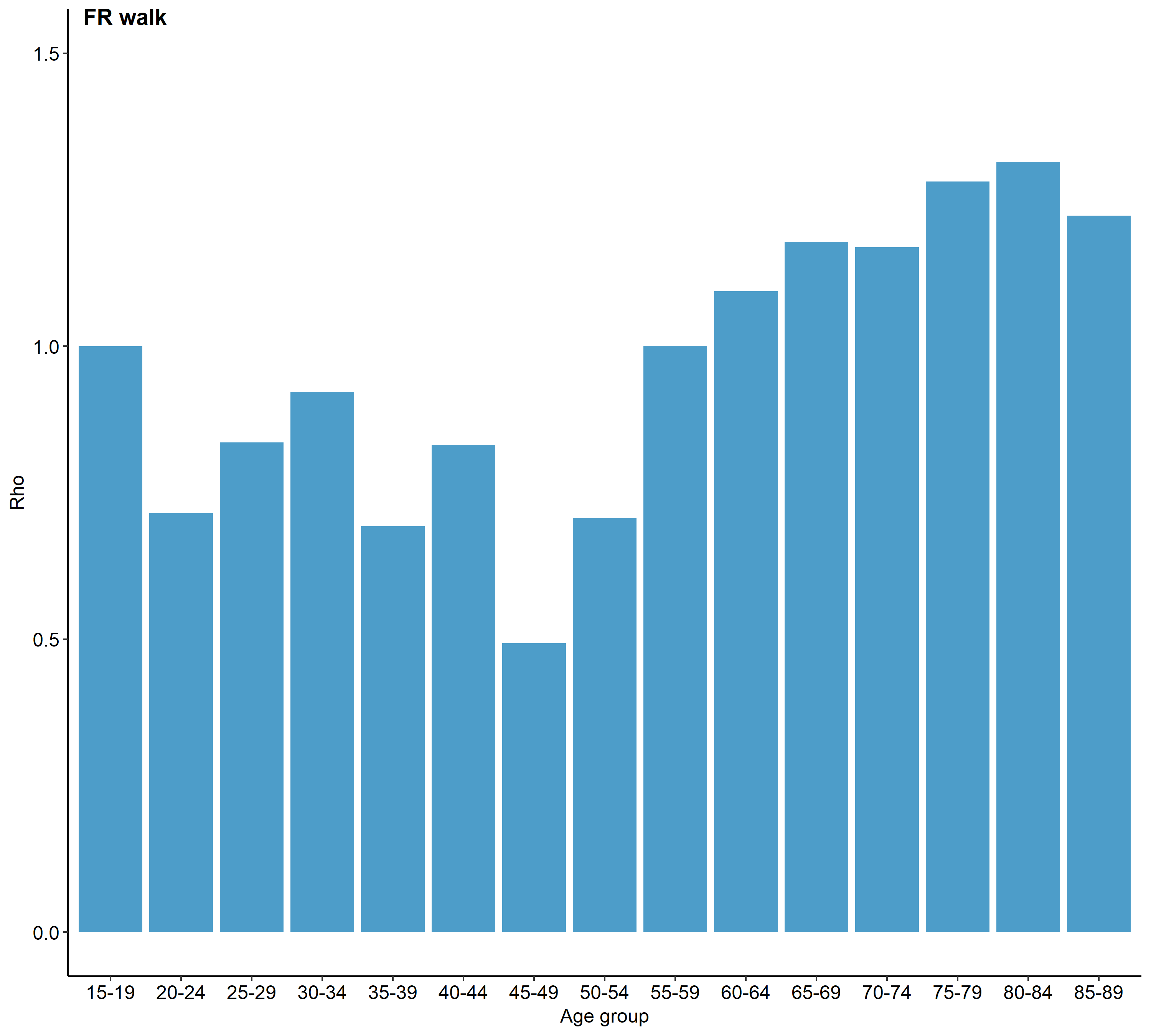

**Figure S4: Age distribution of walking distances across age groups (from 15-19 to 85-89 years) in France**

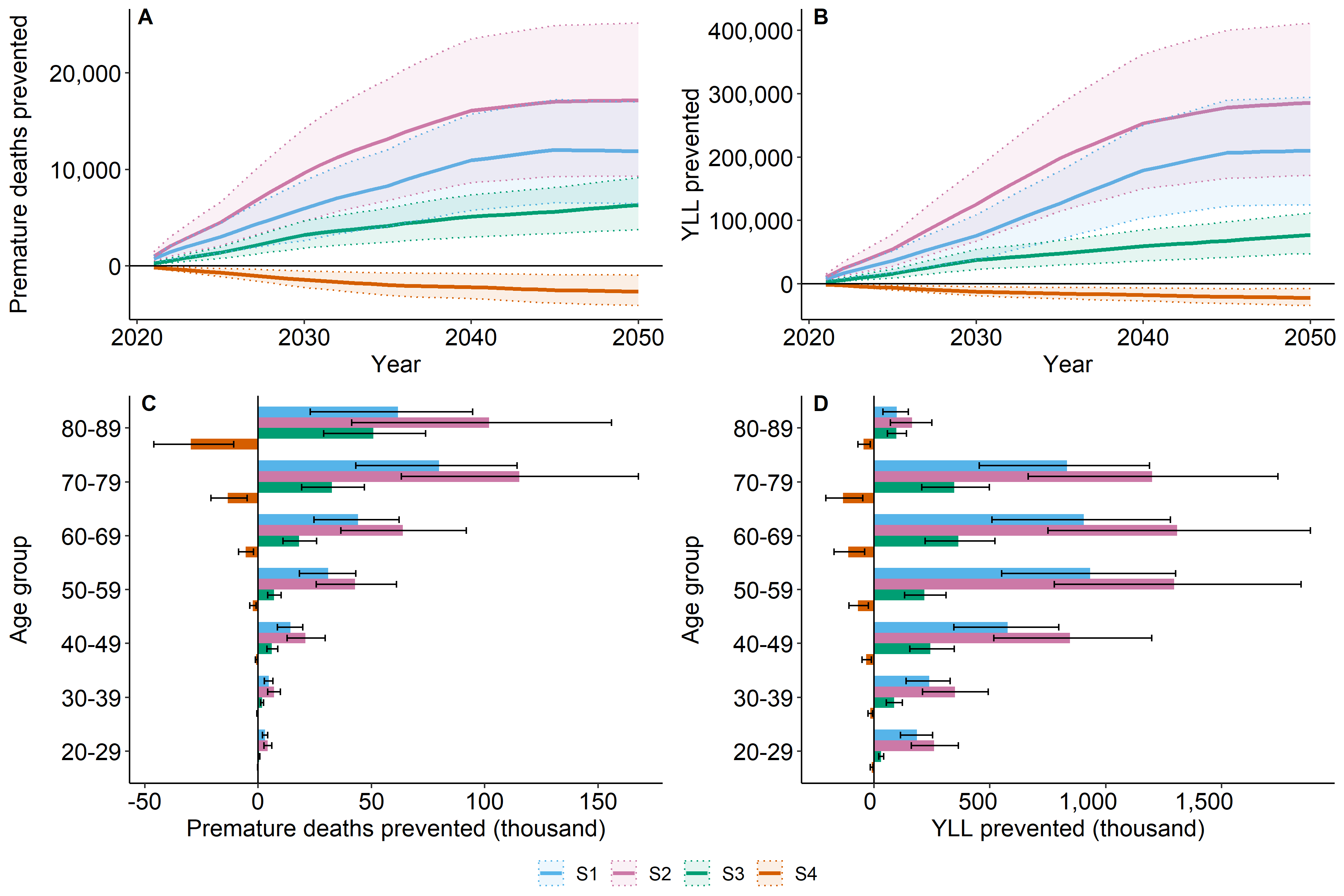

**Figure S5 (sensitivity analyse fourth: evolution of relative contributions for S1 and S2 scenarios) : Deaths prevented (A and C) and YLL prevented (B and D) by year (A and B) and age group (C and D) for each transition scenario compared to the BAU.**

Center for Transport Analytics, Transport, n.d. The Danish National Travel Survey.

Ministry of Ecological Transition, n.d. Enquête mobilité des personnes de 2019.

The French Agency for Ecological Transition, n.d. Transitions 2050 - Portail de données [WWW Document]. URL https://data-transitions2050.ademe.fr/datasets?topics=vKftP1ZR0-SvQtrX4J5pS (accessed 7.26.23).

Zhao, Y., Hu, F., Feng, Y., Yang, X., Li, Y., Guo, C., Li, Q., Tian, G., Qie, R., Han, M., Huang, S., Wu, X., Zhang, Y., Wu, Y., Liu, D., Zhang, D., Cheng, C., Zhang, M., Yang, Y., Shi, X., Lu, J., Hu, D., 2021. Association of Cycling with Risk of All-Cause and Cardiovascular Disease Mortality: A Systematic Review and Dose-Response Meta-analysis of Prospective Cohort Studies. Sports Med. Auckl. NZ 51, 1439–1448. https://doi.org/10.1007/s40279-021-01452-7
